# Phonological Processing is Below Expectations and Linked to Word-Finding Difficulty in Multiple Sclerosis

**DOI:** 10.1101/2023.09.05.23295080

**Authors:** James F. Sumowski, Emily Dvorak, Sarah Levy, Jordyn R. Anderson

## Abstract

**Background:** Word-finding difficulty is prevalent but poorly understood in persons with relapsing-remitting multiple sclerosis (RRMS).

**Objective:** Investigate our hypothesis that phonological processing ability is below expectations and related to word-finding difficulty in patients with RRMS.

**Method:** Data were analyzed from patients with RRMS (n=50) on patient-reported word-finding difficulty (PW-WFD) and objective performance on WIAT-4 Phonemic Proficiency (PP; analysis of phonemes within words), Word-Reading (WR, proxy of premorbid literacy and verbal ability), and Sentence Repetition (SR; auditory processing of word-level information).

**Results:** Performance (mean [95% CI]) was reliably lower than normative expectations for PP (− 0.41[-0.69, -0.13]) but not for WR (0.02[-0.21, 0.25]) or SR (0.08[-0.15, 0.31]. Within-subjects performance was worse on PP than both WR (t[49]=4.00, p<0.001, d=0.47) and SR (t[49]=3.76, p<0.001, d=0.54). Worse PR-WFD was specifically related to lower PP (F_2,47_=6.24, p=0.004, η^2^=0.21); worse PP performance at PR-WFD Often (n=13; -1.16[-1.49, -0.83]) than Sometimes (n=17; -0.14[-0.68, 0.41]) or Rarely (n=20; -0.16[-0.58, 0.27]. PR-WFD was unrelated to WR or SR (Ps>0.25).

**Conclusion:** Phonological processing was below expectations and specifically linked to word-finding difficulty in RRMS. Findings are consistent with early disease-related cortical changes within the posterior superior temporal / supramarginal region. Results inform our developing model of MS-related word-finding difficulty.

## INTRODUCTION

Word-finding difficulty is among the most prevalent cognitive complaints in relapsing-remitting multiple sclerosis (RRMS),^1^ which often leads to embarrassment and frustration in social and employment settings. Patients describe a ‘tip of the tongue’ phenomenon in which they know what they want to say, but cannot retrieve the target word. Despite this, there has been very little research on MS word-finding difficulty, and it remains poorly understood. Neuroimaging research identifies temporal-parietal regions as particularly vulnerable to early cortical atrophy^2-3^ and cortical lesions^4^ in MS, especially the posterior superior temporal / supramarginal region, which is strongly implicated in phonological processing and retrieval of phonemic codes.^5-6^ We investigated whether persons with RRMS have difficulty with phonological processing, and whether phonological processing relates to reported word-finding difficulty.

## METHODS

### Sample

Data were captured through retrospective chart review of our MS cognitive screening program; the IRB of the Icahn School of Medicine at Mount Sinai gave ethical approval for this work. Data were analyzed for consecutive RRMS patients aged 18 to 59 years without other neurologic conditions evaluated during a 2.5-month period when the cognitive battery assessed phonological processing (see below). Data were excluded from persons with the following histories associated with developmental weaknesses in English-language phonemic processing: developmental speech / language impairment, dyslexia, hearing impairment, English learned as a second language after one year of age.^7-9^

### Language Tasks

Patients completed subtests of the Wechsler Individual Achievement Battery, Fourth Edition (WIAT-4,^10^ standardized norm-referenced battery; raw scores converted to age-adjusted normative z-scores). ***Word Reading (WR):*** persons read increasingly sophisticated (lower frequency) individual words aloud; serves as a proxy of premorbid literacy and verbal ability, and also a control for undiagnosed reading weakness. ***Phonemic Proficiency (PP)*:** persons aurally presented with target words and asked to manipulate phonemes to produce new words, including manipulation of sounds within words (e.g., “Say *collect* but change the /l/ sound to /r/.” *correct*), and pronouncing words backwards (e.g., “Say *knife* backwards.” *fine*). Persons receive two points for correct responses within two seconds, one point for correct responses after two seconds, zero points for incorrect responses. ***Sentence Repetition (SR):*** persons repeat aurally-presented sentences of increasing length and complexity. Persons receive two points for perfect repetitions, one point for responses with ≤2 errors, zero points for responses with ≥3 errors. SR is a good comparison for PP; both require processing, maintenance, and production of auditory information, but PP requires processing of phonemes within words, whereas SR requires processing of words within sentences.

### Patient-Reported Word-Finding Difficulty (PR-WFD)

Patients were asked how often (never, rarely, sometimes, fairly often, very often) they experience: (a) “having a word ‘on the tip of your tongue’ but with difficulty getting it out,” (b) “having a sense of what you want to say, but having trouble clearly expressing your thoughts,” (c) “accidentally saying the wrong word / misspeaking.” Responses were coded 0 (never) to 4 (very often). Internal consistency was good (Cronbach’s alpha: 0.89). We used mean response to derive PR-WFD groups: Rarely (<2.0), Sometimes (≥2.0 but <3.0), Often (≥3.0).

**Symbol Digit Modalities Test (SDMT)** is the most sensitive test of overall MS cognitive dysfunction;^11^ we used SDMT to characterize our sample relative to age-adjusted normative data,^12^ and as a comparison task representing generalized MS-related cognitive decline.^11^

### Data Availability

All data produced in the present study are available upon reasonable request to the authors.

### Statistical Analyses

Means and 95% confidence intervals (CI) characterized differences in task performance versus the healthy standardization sample. Dependent t-tests assessed within-subject differences between pairs of tasks. If persons with RRMS have a phonological processing weakness, the PP z-score should be reliably less than zero, and PP should be worse than WR and SR. One-way ANOVA assessed differences in test performance across PR-WFD levels.

## RESULTS

### Sample

We identified 69 RRMS patients aged 18-59 without other neurologic conditions. To control for developmental differences in phonological processing, we excluded data from persons with developmental histories of speech / language delay (n=2), dyslexia (n=3), hearing impairment (n=1), or learning English as a second language after one year of age (n=11). We additionally excluded data from persons with WIAT-4 WR z-scores <-1.5 due to possible undiagnosed dyslexia (n-2). Data were analyzed for the remaining 50 RRMS patients (Table 1). Characterizing the sample cognitively, SDMT age-adjusted normative z-score (mean [95% CI]) was below expectations (−0.36 [-0.63, -0.09]); 30% and 14% of patients had SDMT z-scores ≤-1.0 and ≤-1.5, respectively.

**TABLE 1.** Sample Characteristics.

|  |  |
| --- | --- |
| Age, years, mean [sd] | 41.3 [10.7] |
| Sex, female n : male n | 40 : 10 |
| Education, Bachelor Degree, no n : yes n | 12 : 38 |
| Race / Ethnicity [n] |  |
| Hispanic / Latino(a) | 8 |
| Black, Non-Hispanic | 10 |
| White, Non-Hispanic | 32 |
| Years since diagnosis, median [IQR] | 6 [2, 10] |
| Performance z-scores, mean [SD] |  |
| Word Reading | 0.02 [0.81] |
| Sentence Repetition | 0.08 [0.82] |
| Phonemic Proficiency | -0.41 [0.98] |
| SDMT | -0.36 [0.96] |
| SDMT, raw, mean [sd] | 55.3 [11.5] |
| PR-Word-Finding Difficulty, raw, mean [sd] | 2.0 [1.2] |

### Language Tasks

There were no concerns for skewness, kurtosis, or outliers. Performance (mean [95% CI]) was reliably lower than normative expectations for PP (−0.41 [-0.69, -0.13]) but not for WR (0.02 [-0.21, 0.25]) or SR (0.08 [-0.15, 0.31]. Likewise, percentage of patients performing one standard deviation below expectations (z ≤-1.0) was 3x higher for PP (30%) than for (10%) or SR (8%). Within-subjects performance was worse on PP than both WR (t[49]=4.00, p<0.001, d=0.47) and SR (t[49]=3.76, p<0.001, d=0.54; **Fig 1a-b**).

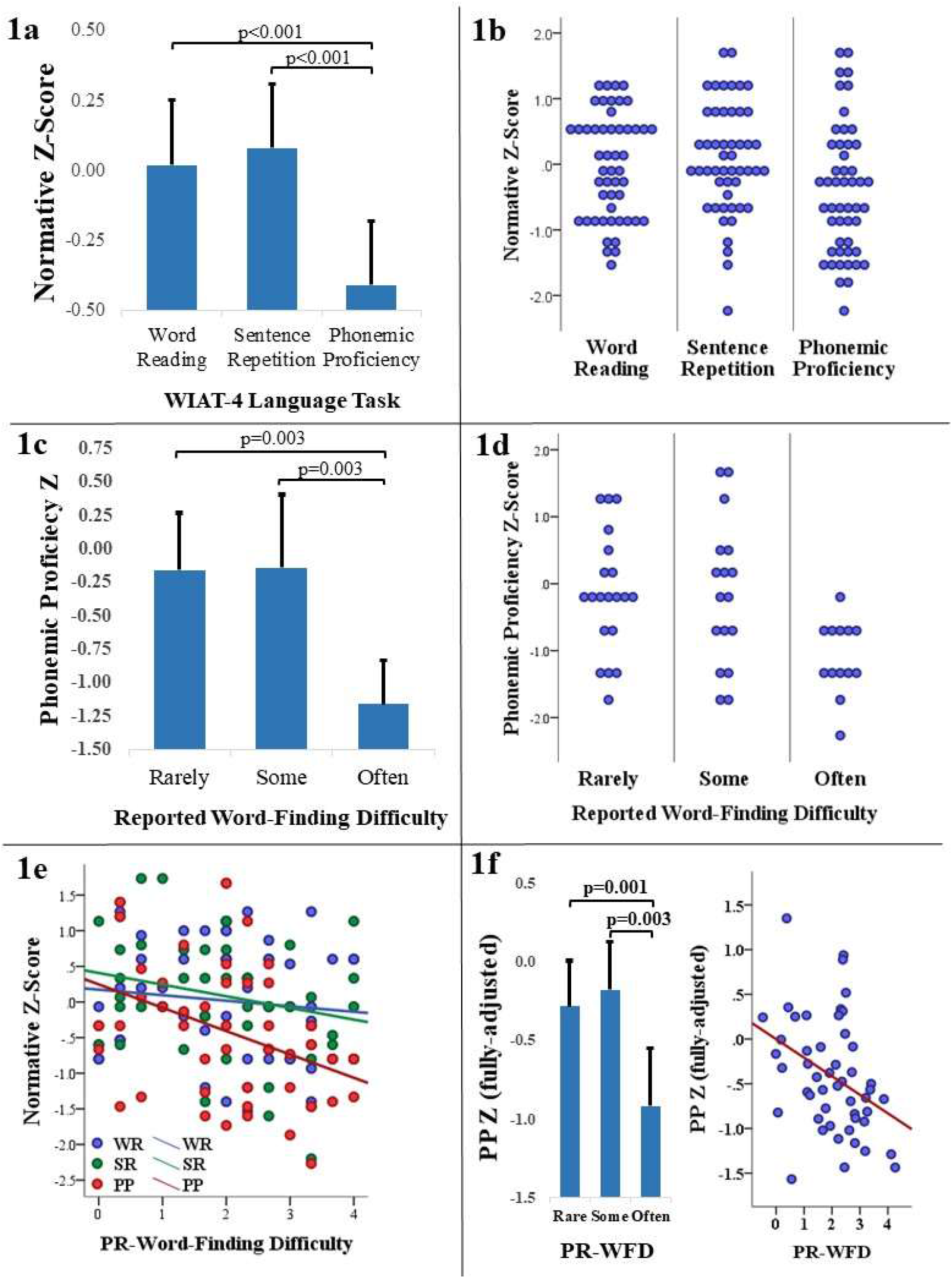
Panels 1a and 1b show differences in age-adjusted normative z-scores across Word Reading (WR), Sentence Repetition (SR), and Phonemic Proficiency (PP), first as means with 95% CIs (1a) and then as a dot plot showing each data point (1b). Panel 1c shows mean PP z-score and 95% CIs across patient-reported word-finding difficulty (PR-WFD) levels (Rarely, Sometimes, Often); panel 1d is a dot plot showing each PP z-score within each PR-WFD level. Panel 1e plots the correlation between PR-WFD (as a continuous variable) and performance on WR (−0.11, p=0.440), SR (−0.24, p=0.101), and PP (−0.39, p=0.005). Panel 1f is based on sensitivity analyses; left side shows means and 95% CIs for PP z-score adjusted for WR, SR, and SDMT across levels of PR-WFD, and the right side of panel 1f provides a scatterplot of the partial correlation between PP z-score and continuous PR-WFD (r_p_= -0.37, p=0.011) adjusting for WR, SR, and SDMT.

### Patient-Reported Word-Finding Difficulty

PR-WFD level was related to PP (F_2,47_=6.24, p=0.004, η^2^=0.21); PP performance was worse at PR-WFD Often (n=13; -1.16 [-1.49, -0.83]) than Sometimes (n=17; -0.14 [-0.68, 0.41]) or Rarely (n=20; -0.16 [-0.58, 0.27], **Fig-1c-d**). PR-WFD was not reliably related to WR (p=0.599) or SR (p=0.299), nor was it related to SDMT as a proxy of general disease-related cognitive decline (p=0.221). Assessed as correlations (**Fig-1e**), worse continuous PR-WFD score was reliably associated with worse PP (r=-0.39, p=0.005) but not WR (r=-0.11, p=0.440), SR (r=-0.24, p=0.101), or SDMT (r=-0.18, p=0.207).

### Sensitivity Analyses

<u>(1) Independent link between PR-WFD and PP:</u> PP was not related to age, sex, education, race/ethnicity, or mood (Hospital Anxiety and Depression Scale; P’s>0.25). Not surprisingly, PP was strongly related to WR (r=0.66), likely due to common neurodevelopmental substrates and the importance of phonological processing for reading. PP was also related to SR (r=0.49; likely due to auditory processing demands) and SDMT (r=0.42; as both appear impacted by disease). ANCOVA reassessed the relationship between PP and PR-WFD adjusting for WR, SR, and SDMT; PP continued to strongly differ across PR-WFD level (F_2,44_=5.32, p=0.009, η^2^=0.20) with worse PP in PR-WFD Often (−0.92 [-1.28, -0.55]) than Sometimes (−0.18 [-0.49, 0.13] or Rarely (−0.28 [-0.57, 0.01]). Likewise, utilizing multiple regression, greater PR-WFD was still predicted by lower PP (r_p_= -0.37, p=0.011; **Fig-1f**) when adjusting for WR, SR, and SDMT (Ps >0.10; VIF’s ≤2.15). <u>(2) Exploring Link to Verbal Memory:</u> Word-finding is a language process rather than a memory function. To be thorough, however, we assessed the link between PR-WFD and word-list learning (Hopkins Verbal Learning Test, Revised; HVLT-R). One-way ANOVA revealed no differences in age-adjusted normative z-scores for HVLT-R Total Learning or Delayed Recall across levels of PR-WFD (P’s >0.50), and no correlations with continuous PR-WFD scores (P’s >0.20). <u>(3) Confirming Normative Deficit:</u> WIAT-4 normative data extend to age 50; we utilized age-50 norms for 14 patients aged 51-59 years. Results remained the same when repeating analyses using data from only patients aged ≤50 years (n=36). Patients still performed worse on PP (−0.34 [-0.63, -0.06]) than on WR (0.07 [-0.19, 0.33]) or SR (0.16 [-0.08, 0.40]), and PP was still the only task that reliably differed across levels of PR-WRD (F_2,33_=4.79, p=0.015, η^2^=0.23; Often [n=6; -1.08 [-1.51, -0.65], Sometimes [n=12; -0.49 [-1.00, 0.02], Rarely [n=18; 0.00 [-0.41, 0.40]).

## DISCUSSION

Persons with RRMS exhibited an objective phonological processing weakness relative to the healthy standardization sample, and relative to their own performance on word reading (proxy of premorbid literacy and verbal ability) and sentence repetition (processing and maintenance of whole-word-level auditory information). Patient-reported word-finding difficulty was specifically associated with weaker phonological processing, which informs our developing model of word-finding difficulty in MS.^1^ A disease-related phonological processing weakness is consistent with neuroimaging evidence of early cortical atrophy^2-3^ and cortical lesions^4^ within the posterior superior temporal / supramarginal region, which is the primary region for phonological processing and phonemic retrieval during word-finding.^5-6^ Research needs to investigate the relationship between word-finding and disease-related changes in this cortical region, and whether this expected relationship is mediated through differential phonological processing ability, which would inform symptomatic treatment development.

## ACKNOWLEDGEMENTS

The authors thank the clinicians, research coordinators, and patients of the Corinne Goldsmith Dickinson Center for MS.

